# Prostad: The development and evaluation of a prostate cancer rapid diagnostic pathway, a protocol

**DOI:** 10.1101/2024.05.30.24308198

**Authors:** Katie Jones, Sarah Rees, Steven Farrington, Arya Chandran, Sohail Moosa, Janet MacKrell, Savita Shanbhag, Yeung Ng, Berni Sewell, Mari Jones, Esra Erdem, Deborah Fitzsimmons, Nick Rich, Jaynie Rance, Rachel Gemine

**Affiliations:** Swansea University; TriTech, Hywel Dda University Health Board; Hywel Dda University Health Board

**Keywords:** rapid diagnosis pathways, prostate cancer, RDCs, cancer diagnosis, health service evaluation

## Abstract

**Background:** Delay to cancer diagnosis is associated with poorer outcomes. In response to delays to cancer diagnosis in the UK, a number of Rapid Diagnosis Centres (RDCs), Multi-Disciplinary Centres (MDCs) and other pathway reforms have been piloted and implemented in recent years. Prostate cancer is the most commonly diagnosed cancer for men or those assigned male at birth in the UK. In Wales, the recommended time to diagnosis is within 62 days from point of suspicion. For patients served by Hywel Dda University Health Board, current waiting times on the prostate cancer diagnostic pathway are prolonged, falling well outside the 28-day decision to treat and 62-day referral to treatment targets. A revised prostate cancer diagnostic pathway called Prostad (Wesh for “prostate”) has been developed and is currently being implemented with the aim of reducing time to diagnosis (or discharge) for patients referred for investigation. This protocol describes Prostad and the planned evaluation approach.

**Methods:** This is a mixed-method evaluation. It is shaped by patient and public involvement throughout and incorporates realist interviews with multiple stakeholders (including NHS staff and patients), process mapping, economic evaluation, and monitoring of the intervention against its aims using routinely collected data.

**Discussion:** To the authors’ knowledge, this is the first project of its kind to combine service aims evaluation, cost-effectiveness analysis and realist evaluation approaches, and as such promises findings applicable to organisations and individuals with regard to various aims and priorities. Continued patient and public involvement throughout the study constitutes one of its strengths.

## Introduction

Delays in the diagnosis of cancer are associated with poorer outcomes,^1^ with earlier diagnosis and detection constituting an important factor in reducing chance of metastases and increasing possibility of curative treatment.^2^ Cancer Research UK (CRUK) found that the survival rate for cancer in the UK is lower than that of comparable countries, and that this is partly due to delays to diagnosis.^3^ Research using data from 2014’s English National Cancer Diagnosis Audit found that 24% of patients experienced avoidable delays to diagnosis, with most of these delays occurring within primary (49%) or secondary (38%) care, as opposed to the pre-consultation (13%) stage.^4^

Responding to these issues, a number of Rapid Diagnosis Centres (RDCs), Multi-Disciplinary Centres (MDCs) and other pathway reforms have been piloted and implemented in recent years, many under the Accelerate, Co-ordinate, Evaluate (ACE) programme.^5^ NHS England’s National Cancer Programme focuses on accelerated diagnosis pathways to ensure patients with suspected cancer receive diagnostic tests to confirm or refute their suspicion within 28 days of referral.^6^ Based on a Danish model of care that responds to similar delays in cancer diagnosis, RDCs aim to expedite diagnosis, particularly for those with vague symptoms.^7^

There is positive evidence for the RDC-style model, broadly conceived. Manchester’s RAPID programme for Lung Cancer, launched in 2016, has successfully addressed delays at the front end of suspected lung cancer pathway, through workforce reorganisation.^8^ Likewise, rapid access one-stop prostate clinics have been shown to shorten the diagnostic pathway.^9 10 11^Sundi *et al*. found that clinical assessment followed by same-day evaluation of prostate cancer patients’ imaging and biopsy results, by a Multi-Disciplinary Team (MDT), led to critical changes in management plans for one in four patients.^12^ Importantly, rapid diagnostic pathways also have potential benefits with regard to cost-effectiveness.^13^

### Prostate Cancer Diagnosis

Prostate cancer is the most commonly diagnosed male cancer in the UK. Data from the National Prostate Cancer Audit 2020 shows a 23% rise in annual prostate cancer diagnoses from 2017.^14^The Welsh Cancer Intelligence Surveillance Unit data shows that across Wales, 3,260 men were diagnosed with prostate cancer in 2018. The number of prostate cancer cases dropped to 2261 in 2020 and 2161 in 2021.^15 16^ However, this data pertains to the first years of mandated quarantine and isolation periods in response to the Covid-19 pandemic and so the reasons for the decrease are unclear. Of the 3260 PCa diagnosis in Wales in 2018, 461 patients (14.2%) were based in west Wales under Hywel Dda University Health Board (HDdUHB), even though the health board represents only 10% of Wales’ population.^16^

### Rationale for Pathway Revision

NICE guidelines for prostate cancer diagnosis recommend a full multi-parametric MRI scan including contrast enhanced imaging and Prostate Cancer UK have supported this by publishing an imaging technical guidance document.^17 18^ Wales’ National Optimal Pathway for prostate cancer aligns with NICE and Prostate Cancer UK.^19^ It describes good practice diagnostic and treatment pathways, stating that the diagnostic pathway, including staging, should take no more than 28 days, with Magnetic Resonance Imaging (MRI) recommended within 7 days and biopsy by Day 14. Organisational reforms and increasing demands on the service mean that the challenges of meeting these cancer targets are exacerbated by a lack of capacity and resources.^20^ State-funded healthcare systems like the NHS are facing unprecedented demand to meet cancer targets following the Covid-19 pandemic.^21^

For patients served by HDdUHB, current waiting times on the prostate cancer diagnostic pathway are prolonged, falling well outside the 28-day decision to treat and 62-day referral to treatment targets. StatsWales indicates that in January 2023, 16.7% of patients covered by HDdUHB began their first definitive treatment within the recommended 62 days of first being suspected of cancer.^22^ This figure is representative of previous and later months, and illustrates the extent to which figures for HDdUHB fall below the Welsh government target of 75%. Urologists and other departments within the health board conducted extensive process-mapping (internal; unpublished) to explore factors contributing to delays, and identify deficiencies in the pathway (versus the optimal national pathway) that related to initial communications with the patient, capacity to offer and report on MRIs, capacity within pathology, outpatient clinic waits both in HDdUHB and Swansea Bay University Health Board.

Based on this process mapping and the optimal national pathway, a new prostate cancer rapid diagnosis pathway, known as Prostad (Welsh for “prostate”), has been developed. It is supported by CRUK “Test, Evidence, Transition” (TET) programme and is currently being trialled in HDdUHB. The characteristics of the conventional pathway are shown in *Figure 1. Conventional Pathway*. For comparison, *Figure 2. Prostad Pathway* illustrates the new pathway for prostate cancer diagnosis. Patients on the new pathway undergo MRI investigation and attend (usually virtually) a consultation where they receive their results the following day. If the recommendation is that a biopsy is required, the aim is to perform a transperineal biopsy within 7 days of receiving the MRI result. This approach requires protected time with the MRI scanner on a specified day, ideally in the morning with reporting taking place in the afternoon.

**Figure 1.**
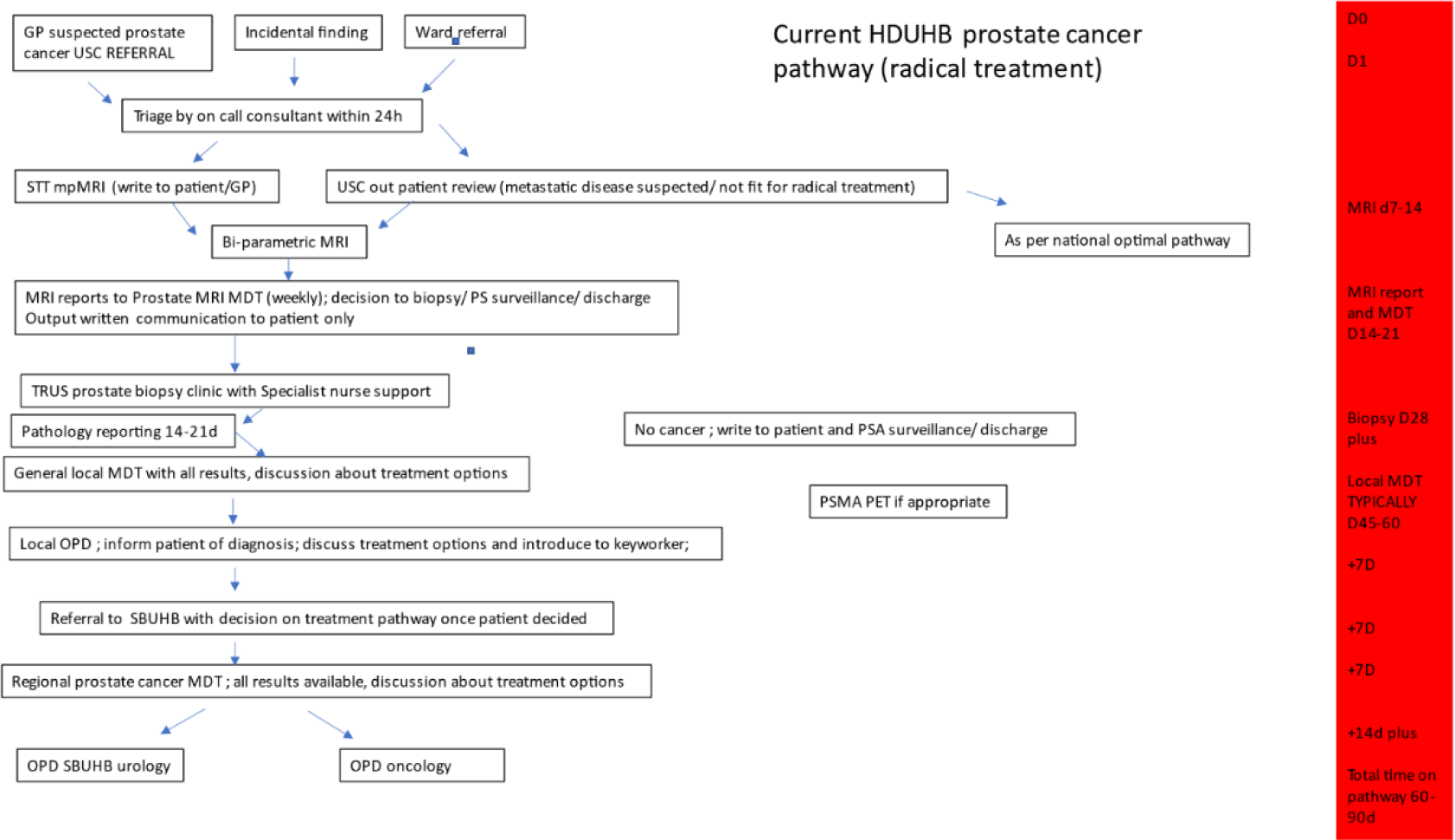
Conventional Pathway.

**Figure 2.**
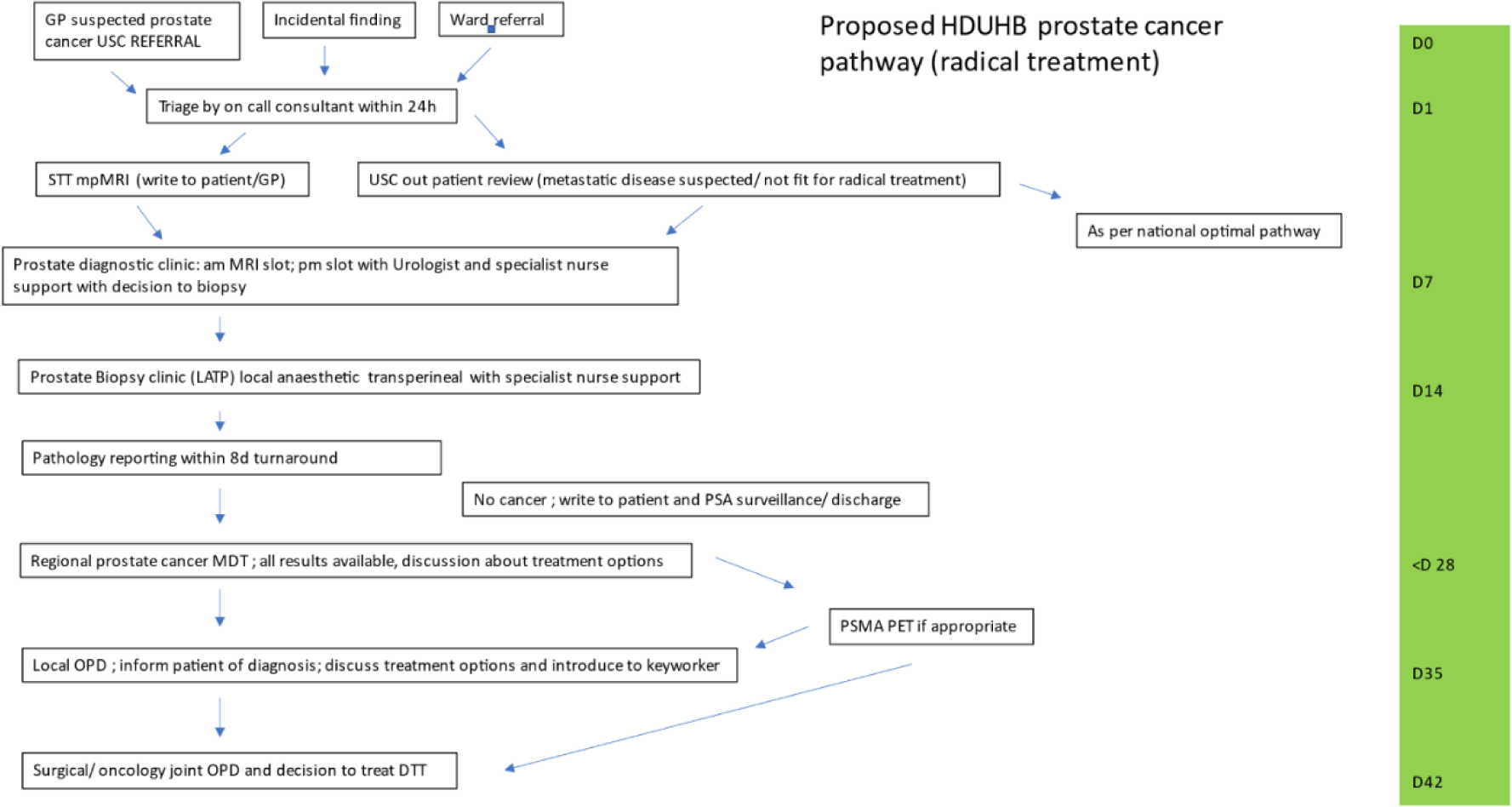
Prostad Pathway.

### Study Overview

This project explores the implementation and impact of the Prostad pathway, against its intended outcomes, such as decreasing waiting times for prostate cancer diagnosis, and with reduced time to diagnosis, earlier treatment. Another aim of the pathway is reduced anxiety for patients who are found to be cancer-free; the theory underpinning this aim is that the time between investigation and result can be stressful and so providing reassurance earlier will be better for patients’ mental health in instances where an MRI and / or biopsy rules out prostate cancer. There is also evidence to indicate that the Prostad pathway will cost less than the conventional pathway.

The aims of this evaluation are:

1. to track, evaluate and explore the process of the Prostad pathway’s development, implementation and delivery;
2. to understand effectiveness based on the intended outcomes (e.g. cost-effectiveness, reduced waiting times);
3. to explore how, for whom and under which circumstances the Prostad pathway produces intended and unintended outcomes;
4. to identify the mechanisms by which the Prostad pathway produces intended and unintended outcomes.

## Materials and Methods

This is a service evaluation with a mixed- and multi-method approach to exploring the implementation of Prostad, intended as a service improvement to the prostate cancer diagnosis pathway. The setting is HDdUHB, a rural area in West Wales that serves a population of around 385,600 inhabitants with 25% of inhabitants being 65 years of age.^23^

There are five arms to this study:

- Work package 1, Patient and Public Involvement (PPI)
- Work package 2, Realist Evaluation
- Work package 3, Service Aims
- Work package 4, Economic Evaluation
- Work package 5, Development of Guidance for the Implementation of Similar Pathways

The project was initiated in March 2023 and is planned to run until August 2024. Prostad, the prostate cancer diagnostic pathway, is currently in its piloting stage and as of January 2024, around 70 patients have utilized the pathway; as a service improvement, consent for referral is not required. It’s worth noting that the conventional pathway remains operational. The evaluation team at Swansea University are conducting a secondary analysis based on data collected by the health board; the health board will obtain explicit consent from patients or other stakeholders who agree to be interviewed. At the time of writing (February 2024), no data has been received for analysis. While all work packages focus on the delivery of the Prostad pathway within the specific context of HDdUHB, it will also consider how the findings can benefit and inform other organisations, including health boards/Trusts and wider stakeholders such as CRUK; this element constitutes work package five (described below).

### Eligibility Criteria and Subject Selection

We will include patients living within the HDdUHB area, who have been referred to Prostad.

### Duration and Timescales

The service evaluation will run over 12 months from August 2023, allowing a 3-month set up phase and 3-month analysis and write up phase. Based on HDdUHB data, we anticipate four patients every week will need an MRI. In line with this, four dedicated MRI slots a week will be available, allowing up to 208 patients to pass through the pathway during the pilot.

### Funding, Ethics and PPI

The project is funded by CRUK and is part of the “Test, Evidence, Transition” programme. It has been informed and supported by collaboration with the West Wales Prostate Support Group. The CRUK funding includes a defined PPI work package to support co-production and a culture of patient inclusion. The evaluation has been approved by the Research, Innovation and Value Based Health Care Department at HDdUHB; NHS research ethics approval was not required for this health board-led evaluation. Swansea University Ethics Committee granted its favourable opinion on 9^th^ January 2024.

### Work package 1, Patient and Public Involvement (PPI)

In December 2023, a PPI group with experience of prostate cancer diagnosis was established. They will inform various elements of the project until its completion, currently planned for August 2024. Members of the PPI group have provided feedback on patient-facing literature and documentation relating to the Prostad pathway (e.g. letters of referral; a leaflet about the service).

The group have been or will be consulted regarding:

- materials and literature aimed at patients referred to Prostad;
- question formulation and refinement of initial theories;
- findings and analysis of qualitative data (April/ May 2024); and
- dissemination of our findings (July/ August 2024).

### Work package 2, Realist Evaluation

Realist evaluation^24^ is a widely used mixed-methods approach which aims to generate theories regarding the relationship between the context (C) in which an intervention is placed and the mechanisms (M) by which an intervention produces outcomes (O). A realist approach holds that, while the intervention may remain the same, contextual factors and the way people interact with an intervention influence the mechanisms by which it may work for some people. Particularly useful when considering the introduction of multifaceted interventions in complex environments, realist evaluation holds that an intervention in and of itself does not necessarily create change, but rather it is how individuals interact with and respond to an intervention that promotes (or fails to promote) change.^25^ We will use a realist approach to produce programme theories relating to *how, for whom, and under which circumstances Prostad works or fails to work* as measured against intended *and* unintended outcomes.

More specifically, this work package aims to:

1. explore the process of implementing the new prostate cancer diagnostic pathway, the Prostad pathway, and develop theories to inform guidance applicable to the implementation of similar pathways elsewhere.
2. identify the outcomes (intended and unintended) of Prostad on multiple stakeholders, including staff and patients;
3. identify the mechanisms by which Prostad produces outcomes for staff and patients;
4. identify the contextual factors that impact the mechanisms by which Prostad produces outcomes for multiple stakeholders.

This work package includes:

- a realist review (Prospero Registration: CRD42024534962);
- realist interviews conducted with patients who have been referred to the pathway, carers and partners of those patients, clinical staff (e.g. urologists, radiographers) and other stakeholders key to the development, implementation and ongoing delivery of the prostate cancer diagnostic pathway.
- the development of theories regarding the contexts and mechanisms by which Prostad produces intended and unintended outcomes.

The realist evaluation will also utilise findings from work packages three and four to inform theory development.

Participants must be aged over 18 years, and either:

- Involved in the design, implementation and delivery of Prostad; or
- Patients and carers of patients who have passed through the pathway during the pilot phase of Prostad.

Realist interview schedules have been developed (JR and AC); interviews will be conducted virtually (phone or Teams) and then transcribed by one researcher based within the health board (AC); secondary realist analysis of the anonymized transcripts will be conducted by researchers at Swansea University (KJ and JR). In line with realist evaluation norms, interview schedules may be adapted iteratively as data are collected.

We are aiming for around 40 realist interviews in total.

### Work package 3, Service Aims Evaluation

This will assess the service delivery against its aims to reduce overall time on pathway, reduce unnecessary activities and improve efficiencies within the service by identifying pinch points and identifying real time solutions. An implementation review will be conducted to consider the barriers and facilitators to implementation. In line with Lean Development,^26^ a real time plan-do-study-act cycle will be adopted with scientific methods applied on a continuous basis to formulate a plan, implement the plan, and analyse and interpret the results, followed by development of any required changes.^27^ This will be conducted through a mixed-methods approach, providing a practical way to understand the multiple perspectives, causal pathways, and multiple types of outcomes and to compare current state and future state of the process including safety, staff morale, quality, delivery dependability and costs. Focusing on the pathway, this work package will support optimisation. Discussions with staff involved in the pathway will aid understanding of barriers and facilitators to implementation. Continuous data review will be undertaken to identify pinch points and modify the pathway to make as efficient as possible. An implementation plan will be constructed to focus on outcomes including acceptability to patients, staff and clinical services, wider adoption, appropriateness and feasibility of the pathway (linked to work package 2), fidelity and applicability outside of the evaluation and to other organisations, implementation costs and sustainability in the medium and long-term and in the event of critical disruption.

### Work package 4, Economic Evaluation

The economic evaluation will consider resource use and cost differences between the pilot pathway and current pathways (based on historic matched controls) and patient outcomes (using data obtained from study records and relevant literature) as part of a cost-consequences analysis. This element of the evaluation utilizes data routinely collected by the health board in relation to the service; all data is fully anonymized prior to its transfer to Swansea University staff for analysis (BS; MJ; EE; DF).

Within this work package, a health economic service evaluation of the pilot pathway compared to the current pathway will be undertaken, using resource use and cost data, literature-derived inputs and Patient Reported Experience Measures (PREM)/ Patient Reported Outcome Measures (PROM) data (where available). Potential costs and consequences (including health outcomes should data be available) of introducing the Prostad pathway and whether this could be considered value for money for the health board will be explored. As part of this work package, a health economic analysis plan, detailing the data to be collected will be developed and agreed with the HDdUHB team and CRUK prior to the analysis.

Specific objectives of the health economic evaluation will be to map out the Prostad pathway in an agreed specific patient population, to understand the impact of the service when compared to ‘standard clinical practice’ (i.e., with no Prostad pathway) on key descriptives such as referrals patterns and time to event across the diagnosis pathway; to identify key resource drivers and costs associated with the Prostad pathway service and subsequent impact on other NHS resources; to investigate the impact of the Prostad pathway on for example, cancers detected, stage of diagnoses; to assess short-term outcomes for patients and to explore the cost-consequences of the Prostad pathway (should data allow) in improving outcomes.

The following PICO will guide the health economic evaluation:

| Population | Intervention | Comparison | Outcomes |
| --- | --- | --- | --- |
| Men with suspicion of prostate cancer referred by their GP or consultant to the prostate cancer diagnosis services<br>• Intervention group: men going through the new pathway<br>• Comparator group: men who have gone through the standard pathway<br>No subgroups will be analysed. | Prostad - Model<br>Prostate Cancer<br>Diagnostic Pathway | Standard pathway in<br>HDdUHB (base case) | 1. Time to diagnosis<br>2. Cancers detected<br>3. Other significant diagnoses<br>4. Anxiety and depression scores<br>5. Health-related quality of life<br>6. Pathway costs<br>7. Healthcare resource use between referral and diagnosis<br>8. Patient experience and satisfaction |

As the new pathway undergoes continuous development, we will work with all relevant stakeholders to clearly define the new pathway, and update the patient population, relevant comparator and outcomes of interest (the PICO) as appropriate.

The health economic service evaluation will be undertaken in five steps to address the health economic objectives:

#### 1. Mapping out the Prostad pathway

Discussions with the clinical and study teams, and data collected as part of work packages 1, 2 and 3, will aid mapping of the patient journey on the Prostad pathway and the standard pathway in HDdUHB. This will include a graphic representation of the different stages of the patient journey (e.g., outpatient appointment, USC, MRI, MDT) and timings of the different stages, taken over the course of a selection of clinics by AC. Pathway maps will be reviewed and signed off by the project management group (including PPI representatives) before they are used to develop a patient simulation model aimed at comparing costs and outcomes (until diagnosis) of the two different diagnostic pathways.

#### 2. Identifying the key resource drivers and costs associated with the Prostad pathway service and subsequent impact on other NHS resources

Resource use and costs will be assessed from a UK NHS perspective with costs expressed in 2023/24 £ sterling. No discounting will be applied as the model time horizon does not exceed one year. The cost of the Prostad pathway service (including oncosts and overheads) will be sourced from the HDdUHB finance department and supplemented by discussions with the project and clinical team where required. Local costs will be used where possible to reflect the local scope of the evaluation. Where no local costs are available, healthcare resource use for intervention and comparator patients will be valued using published unit costs with older costs inflated using relevant price indices (if required). The impact of using national standard unit costs will be examined during sensitivity analysis. Use of healthcare resources, including outpatient appointments, inpatient admissions, diagnostic tests and imaging, will be collected through retrospective review of patient data by AC.

#### 3. To investigate the impact of the Prostad pathway, for example, on cancers detected, stage of diagnoses (if available)

Patients going through diagnostic services in either the new or existing pathway, will be divided into different outcome groups, depending on the clinical outcome at diagnosis, including cancer detected, other significant diagnosis, further investigation required and discharge back to GP. These outcomes will be collected through retrospective review of patient files and service notes by AC for both the intervention and comparator groups. If possible and available, cancer stage at diagnosis will also be recorded for both groups.

#### 4. To assess the short-term outcomes for patients

In addition to clinical outcome at diagnosis, short-term outcomes for patients will include:

- time from referral to diagnosis obtained from patient records and service files.
- patient quality of life/utility as assessed using the EuroQol EQ-5D-5L questionnaire routinely collected as part of the diagnostic pathway.
- anxiety experienced as the patient goes through the pathways using the Hospital Anxiety and Depression Scale (HADS) routinely collected as part of the diagnostic pathway.
- patient experience and satisfaction using the National Strategic Clinical Network for Cancer’s Patient Experience Measure (PREM) routinely collected as part of the diagnostic pathway.

Outcomes will be collected and recorded as part of the routine service delivery to enable service evaluation and continuous improvement. Anonymised data will be shared with the Swansea University team enable the health economic service evaluation.

#### 5. To explore the cost-consequences of the Prostad pathway (should data allow) in improving outcomes for people

An economic model will be developed, to estimate the costs and consequences of the Prostad pathway compared to the standard pathway. We expect that a *de novo* model will need to be constructed. Based on data availability, an appropriate model type and structure will be developed to reflect the patient pathway, ensuring that all relevant aspects are captured (including ‘downstream’ consequences of initial decisions) to the point where the ‘assessment’ of effectiveness is agreed. Due to time and budget constraints and the novelty of the pathway, it is not likely that a full ‘life-time horizon’ will be considered in our model, but it will focus on the shorter-term impact of the Prostad pathway on cancer or other diagnoses detected, based on achieving a timelier diagnosis that would be deemed of high value to HDdUHB.

The model will be informed by the implementation costs for the Prostad pathway, healthcare costs for both comparator pathways and outcomes collected in previous steps of the health economic service evaluation. To avoid over-complexity, aggregate costs may be appropriate when trying to capture the overall cost associated with downstream events. Where local data sources are insufficient, unavailable or unfeasible for collection by HDdUHB, the literature or UK sources will be used to identify suitable data inputs. Where this cannot be obtained, appropriate assumptions will be made in conjunction with the HDdUHB project team. All inputs will be agreed prior to analysis and the sources of information will be fully referenced. A descriptive summary of the resources and costs associated with the Prostad pathway compared to the standard pathway will be provided. The costs of the pilot pathway and comparator will then be compared to relevant outcomes as part of a cost-consequences analysis. If data availability allows, we will also undertake an exploratory cost-utility analysis using quality-adjusted life years (QALYs) obtained from EQ-5D-5L responses.

### Sensitivity analyses

Probabilistic and deterministic sensitivity analyses will be undertaken to estimate the uncertainty around results. Scenario analyses will be agreed and undertaken with the HDdUHB team to address ‘What If?’ questions based on the impact of changing key parameters within the pathway on patient outcomes and waiting times and time within the pathway. Based on data availability, scenarios including longer-term extrapolations and agreed upon in advance with HDdUHB and CRUK, may be considered.

### Work package 5, Developing Implementation and Service Guidance

This work package depends and relies upon the outcomes of work packaged 1-4; no additional participants will be recruited for this element of the project. Working with health board staff (SS, SF and AC), the implementation team will support the HDdUHB Urology team and the National Strategic Clinical Network for Cancer in the development of implementation and service guides for national roll-out utilising the TIDieR framework (Hoffman et al, 2014). The guidance will be informed by work packages 1-4 and will include information regarding facilitators and barriers to change in addition to contextual and mechanistic factors that produced intended and unintended outcomes related to the implementation of Prostad. This guidance will aim to enable other clinical teams to understand if, whether and how to make similar service changes to their prostate cancer diagnostic service.

## Discussion

Urologists and general practitioners in collaboration with CRUK have led the development of Prostad, which can be considered alongside the growing number of rapid diagnostic pathways that respond to the problem of long waiting times and NHS priorities.^28 29^ Prostad aims to reduce the time to diagnosis for people with prostate cancer. The multi-and mixed-method approach reflects the intention of this evaluation, namely, to produce a series of outputs that will be relevant to varied stakeholders, including patients, health professionals locally and globally designing and implementing similar centres, and to policy-makers. Evaluation outputs include:

- Project report capturing key TET objectives and economic evaluation based on:

o Time to diagnosis
o Cancer detected
o Other significant diagnoses
o Anxiety and depression scores
o Health-related quality of life
o Pathway costs
o Healthcare resource use between referral and diagnosis
o Patient experience and satisfaction
o Patient and clinician feedback
- Pathway specific documentation
- Training and service planning guides
- Business case for roll-out
- Implementation handbook
- Generalisable route to scale guide
- Summaries for engaging a variety of stakeholders using different media.

To the authors’ knowledge, this is the first project of its kind to combine service aims evaluation, cost-effectiveness analysis and realist evaluation approaches, and as such promises findings applicable to organisations and individuals with regard to various aims and priorities. Continued PPI throughout the study constitutes one of its strengths.

### Limitations of the Evaluation design

While the pathway spans multiple sites, as an innovation/pilot study, this evaluation focuses on a single rapid diagnosis pathway in West Wales. We aim to account for and differentiate between contextual factors that may be singularly or broadly applicable via the realist evaluation methodology.

### Dissemination plans

Our PPI group will be integral to shaping elements of our dissemination plan. However, currently, we plan to host a patient and clinician information event in partnership with the National Strategic Clinical Network for Cancer utilising the urology cancer site group, to discuss and showcase the service to others. In addition, we will disseminate the outcomes through professional channels in urology and quality/patient safety e.g. presentations at BAUS (British Association of Urological Surgeons), NHS Patient Safety Conference, International Forum on Quality and Safety, and publications in relevant journals.

### Amendments

Revisions to this study will be reported to CRUK and tracked and reported in our monthly project meetings. Significant changes to the service during implementation and piloting will be captured as part of the evaluation process and reported through publications. With agreement from the publisher, amendments may also be reported by protocol addendum.

## Authors’ contributions

RG, YN, SM, SR, SS, KJ, SF and AC all contributed to the introduction and pathway description; JR and KJ designed and described the evaluation approach for WP1 and WP2; NR conceptualised the procedure and composed the description of WP3; BS, MJ, and EE designed and described WP4; SS designed and authored WP5. KJ edited the contributions and redrafted them into a format aligning with the journal requirements; all named authors provided editorial input, approved the submitted version, and take responsibility for its accuracy.

## Funding

This project is funded by Cancer Research UK and is part of the “Test, Evidence, Transition” programme.

## Competing Interests

There are no competing interests to declare.

## Data Availability

No datasets were generated or analysed during the current study. All relevant data from this study will be made available upon study completion.

## Notes

### Competing Interest Statement

The authors have declared no competing interest.

### Funding Statement

Yes

### Author Declarations

The evaluation has been approved by the Research, Innovation and Value Based Health Care Department at Hywel Dda University Health Board NHS research ethics approval was not required for this health board-led evaluation. Swansea University Ethics Committee granted its favourable opinion on 9th January 2024.

## References

1 Neal RD, Tharmanathan P, France B, Din NU, Cotton S, Fallon-Ferguson J, Hamilton W, Hendry A, Hendry M, Lewis R, Macleod U, Mitchell ED, Pickett M, Rai T, Shaw K, Stuart N, Tørring ML, Wilkinson C, Williams B, Williams N, Emery J. Is increased time to diagnosis and treatment in symptomatic cancer associated with poorer outcomes? Systematic review. Br J Cancer. 2015 Mar 31;112 Suppl 1(Suppl 1):S92–107. doi: 10.1038/bjc.2015.48.

2 Tobore T.O. On the need for the development of a cancer early detection, diagnostic, prognosis, and treatment response system. Future Sci OA. 2019 Nov 29;6(2): FSO439. doi: 10.2144/fsoa-2019-0028.

3 CRUK. Cancer in the UK. *CRUK,* 2018: www.cancerresearchuk.org/sites/default/files/state_of_the_nation_apr_2018_v2_0.pdf.

4 Swann R, Lyratzopoulos G, Rubin G, Pickworth E, McPhail S. The frequency, nature and impact of GP-assessed avoidable delays in a population-based cohort of cancer patients. Cancer Epidemiol. 2020 Feb;64:101617. doi: 10.1016/j.canep.2019.101617. Epub 2019 Dec 3. PMID: 31810885.

5 CRUK. Key Messages from the Evaluation of the ACE Programme. 2020: https://www.cancerresearchuk.org/sites/default/files/key_messages_ace_evaluation_july_2020_fv1.0_ext.pdf

6 NHS England Achieving world-class cancer outcomes: Taking the strategy forward. NHS, 2016. www.england.nhs.uk/wp-content/uploads/2016/05/cancer-strategy.pdf

7 Vedsted P, Olesen F. A differentiated approach to referrals from general practice to support early cancer diagnosis - the Danish three-legged strategy. Br J Cancer. 2015 Mar 31;112 Suppl 1(Suppl 1):S65–9. doi: 10.1038/bjc.2015.44. PMID: 25734387; PMCID: PMC4385978.

8 Evison M, Hewitt K, Lyons J, Crosbie P, Balata H, Gee C, Duerden R, Greaves M, Sharman A, Booton R. Implementation and outcomes of the RAPID programme: Addressing the front end of the lung cancer pathway in Manchester. Clin Med (Lond). 2020 Jul;20(4):401–405. doi: 10.7861/clinmed.2019-0218. PMID: 32675147; PMCID: PMC7385808.

9 McCombie SP, Hawks C, Emery JD, Hayne D. A ‘One Stop’ Prostate Clinic for rural and remote men: a report on the first 200 patients. BJU Int. 2015 Oct;116 Suppl 3:11–7. doi: 10.1111/bju.13100. Epub 2015 Jul 27. PMID: 26218767.

10 Kavanagh AG, Lee JC, Donnelly B. Time to treatment of prostate cancer through the Calgary Prostate Institute rapid access clinic. Can J Urol. 2008 Apr;15(2):3975–9. PMID: 18405444.

11 Bolton EM, Kelly BD, Quinlan MR, D’Arcy FT, Azar M, Dowling CM, Power M, McCarthy P, Roche C, Walsh K, Rogers E, Durkan GC.. Audit of rapid access introduction reveals high prevalence of prostate cancer in Western Region. Ir J Med Sci 183, 2014, pp. 173–179 10.1007/s11845-013-0986-y

12 Sundi D, Cohen JE, Cole AP, Neuman BP, Cooper J, Faisal FA, Ross AE, Schaeffer EM. Establishment of a new prostate cancer multidisciplinary clinic: Format and initial experience. Prostate. 2015 Feb;75(2):191–9. doi: 10.1002/pros.22904. Epub 2014 Oct 13. PMID: 25307625; PMCID: PMC4270998.

13 Sewell, B., Jones, M., Gray, H., Wilkes, H., Lloyd-Bennett, C., Beddow, K., Bevan, M., & Fitzsimmons, D. Rapid cancer diagnosis for patients with vague symptoms: a cost-effectiveness study. The British journal of general practice : the journal of the Royal College of General Practitioners, 70(692), 2020 e186–e192. 10.3399/bjgp20X708077

14 NPCA. National Prostate Cancer Audit Annual Report. 2020 https://www.npca.org.uk/content/uploads/2021/01/NPCA-Annual-Report-2020_Final_140121.pdf

15 NPCA. An audit of the care received by people with prostate cancer in England and Wales from 01/01/2019 to 31/01/20. NPCA State of the Nation Report, 2024. Retrieved from: Untitled (npca.org.uk)

16 NHS Wales. Cancer Incidence in Wales 2002-2020. Cancer Intelligence Unit. 2023: https://phw.nhs.wales/services-and-teams/welsh-cancer-intelligence-and-surveillance-unit-wcisu/cancer-reporting-tool-official-statistics/cancer-incidence/

17 NICE. Prostate Cancer Diagnosis and Management. NICE Guidelines, 2019 Recommendations | Prostate cancer: diagnosis and management | Guidance | NICE

18 Prostate Cancer UK. Multiparametric Magnetic Resonance Imaging, Prostate Imaging Guidance. 2018 https://prostatecanceruk.org/media/jobjikpi/mpmri-imaging-guidance-document-final.pdf

19 NHS Wales. National Optimal Pathway for Prostate Cancer (2^nd^ Edition), 2023. Retrieved: <u>Suspected Cancer</u> <u>Pathway - NHS Wales Executive</u>

20 Melby L, Håland E. When time matters: a qualitative study on hospital staff’s strategies for meeting the target times in cancer patient pathways. BMC Health Serv Res. 2021 Mar 9;21(1):210. doi: 10.1186/s12913-021-06224-7. PMID: 33750379; PMCID: PMC7941937.

21 World Health Organisation. Essential health services face continued disruption. WHO, 2022: Essential health services face continued disruption during COVID-19 pandemic (who.int)

22 Stats Wales. Suspected Cancer Pathway, 2024. Retrieved from: Suspected cancer pathway (closed pathways): The number of pathways where the patient started their first definitive treatment and those informed they do not have cancer by local health board, tumour site, age group, sex, measure and month (gov.wales)

23 Stats Wales. Population estimates by local health board and age. 2022: https://statswales.gov.wales/Catalogue/Population-and-Migration/Population/Estimates/Local-Health-Boards/populationestimates-by-lhb-age

24 Ray Pawson, and Nicholas Tilley. Realistic Evaluation. SAGE Publications, 1997.

25 Jagosh, J., Tilley, N., & Stern, E. Realist evaluation at 25: Cumulating knowledge, advancing debates and innovating methods. Evaluation, 2016; 22(3): 267–269. 10.1177/1356389016656502

26 Poppendieck M. Principles of lean thinking. IT Management Select. 2011;18(2011):1–7.

27 Langley, G. J., Moen, R. D., Nolan, K. M., Nolan, T. W., Norman, C. L., & Provost, L. P. The improvement guide: a practical approach to enhancing organizational performance. John Wiley & Sons, 2009.

28 NHS England. Faster Diagnosis Framework, 2019: https://www.england.nhs.uk/publication/cancer-programme-faster-diagnosis-framework/

29 NHS Wales. Rapid Diagnosis Clinics. 2021: https://executive.nhs.wales/functions/networks-and-planning/cancer/workstreams/rapid-diagnosis-clinics-programme/

